# Saturation genome editing of *RNU4-2* reveals distinct dominant and recessive neurodevelopmental disorders

**DOI:** 10.1101/2025.04.08.25325442

**Authors:** Joachim De Jonghe, Hyung Chul Kim, Ayanfeoluwa Adedeji, Elsa Leitão, Ruebena Dawes, Yuyang Chen, Alexander JM Blakes, Cas Simons, Rocio Rius, Javeria R Alvi, Florence Amblard, Christina Austin-Tse, Sarah Baer, Elsa V Balton, Pierre Blanc, Daniel G Calame, Charles Coutton, Chloe A Cunningham, Nitsuh Dargie, Katrina M Dipple, Haowei Du, Salima El Chehadeh, Ian Glass, Joseph G Gleeson, Olivier Grunewald, Paul Gueguen, Radu Harbuz, Marie-Line Jacquemont, Richard J Leventer, Pierre Marijon, Olfa Messaoud, Tipu Sultan, Christel Thauvin, Catherine Vincent-Delorme, Elif Yilmaz Gulec, Julien Thevenon, Rodrigo Mendez, Daniel G MacArthur, Christel Depienne, Caroline Nava, Nicola Whiffin, Gregory M Findlay

## Abstract

Recently, *de novo* variants in an 18 nucleotide region in the centre of *RNU4-2* were shown to cause ReNU syndrome, a syndromic neurodevelopmental disorder (NDD) that is predicted to affect tens of thousands of individuals worldwide^1,2^. *RNU4-2* is a non-protein-coding gene that is transcribed into the U4 small nuclear RNA (snRNA) component of the major spliceosome^3^. ReNU syndrome variants disrupt spliceosome function and alter 5’ splice site selection^1,4^. Here, we performed saturation genome editing (SGE) of *RNU4-2* to identify the functional and clinical impact of variants across the entire gene. The resulting SGE function scores, derived from variants’ effects on cell fitness, discriminate ReNU syndrome variants from those observed in the population and dramatically outperform *in silico* variant effect prediction. Using these data, we redefine the ReNU syndrome critical region at single nucleotide resolution, resolve variant pathogenicity for variants of uncertain significance, and show that SGE function scores delineate variants by phenotypic severity. Further, we identify variants impacting function in regions of *RNU4-2* that are critical for interactions with other spliceosome components. We show that these variants cause a novel recessive NDD that is clinically distinct from ReNU syndrome. Together, this work defines the landscape of variant function across *RNU4-2*, providing critical insights for both diagnosis and therapeutic development.

## INTRODUCTION

The spliceosome is a large ribonucleoprotein (RNP) complex that mediates RNA splicing. *De novo* variants in a gene encoding one of the small nuclear RNA (snRNA) components of the spliceosome, *RNU4-2*, were recently shown to cause ReNU syndrome, a prevalent neurodevelopmental disorder (NDD)^1,2^. ReNU syndrome is a complex multi-system disorder characterised by moderate to severe global developmental delay, intellectual disability, hypotonia, acquired microcephaly, speech and motor difficulties, low bone density, and often seizures^1,4^.

*RNU4-2* encodes the U4 snRNA, which is a critical component of the major spliceosome. In particular, U4 is tightly bound with the U6 snRNA in the U4/U6.U5 tri-snRNP and the U4/U6 duplex needs to be unwound for activation of splicing^3^. Variants identified in individuals with ReNU syndrome cluster in an 18-nucleotide (nt) region in the centre of *RNU4-2* which is depleted of variants in population datasets (the ‘critical region’, or CR)^1^. This region is known to accurately position U6 for recognition of the 5’ splice site. Consistent with this, variants causing ReNU syndrome have been shown to alter 5’ splice site usage^1^, with this disruption correlating with phenotype severity^4^. Similarly, variants in two distinct structures within the 18-nt CR (the T-loop and Stem III) have been proposed to differ in clinical severity^4^.

The precise relationship between genetic variation in *RNU4-2* and clinical impact remains incompletely characterised. The variants initially characterised in individuals with ReNU syndrome are all within the 18-nt CR, however, more recent work has proposed a role for variants outside this region, in the 5’ stem loop^5^. It is unclear which, if any, variants outside of the CR could also cause NDD. This is particularly important as the elevated mutation rate of *RNU4-2* and other snRNA genes means that there will be many chance occurrences of variants amongst sequenced individuals with syndromic NDD^6^. Up to 75% of individuals with ReNU syndrome have the same single nucleotide insertion (n.64_65insT). Whether the high recurrence of this particular variant is due to ascertainment bias, germline selection, and/or an elevated mutation rate is currently unknown. Further, it is unclear if available variant effect predictors (e.g. CADD^7^) can effectively distinguish between pathogenic and benign variants in *RNU4-2*.

Resolving these questions will be critical to ensure accurate, comprehensive diagnoses of individuals affected by ReNU syndrome. One approach to clarifying variant impact is through the generation of functional data of variant effect, which can mechanistically inform why specific variants cause disease and improve clinical interpretation of rare variants^8^. However, no experimental assay has yet been established to evaluate variants in *RNU4-2*, owing to its recent association with NDD.

Saturation genome editing (SGE) is a powerful approach to delineate genotype-phenotype relationships^9^. Crucially, it doesn’t rely on variants being observed in an individual with or without disease. Instead, every possible variant across a gene or region can be engineered and the relative functional effects of each determined through a cellular readout. SGE experiments have been performed across numerous protein coding-disease genes, including *BRCA1^10^*, *CARD11^11^*, *DDX3X^12^*, *VHL^13^*, and *BAP1^14^*. In each case, the SGE assay has accurately differentiated between known pathogenic and benign variants.

Here, we present the first SGE of a human non-coding RNA. We developed a strategy to combat the high sequence homology between *RNU4-2* and its many homologs and pseudogenes, obtaining a variant effect map that effectively distinguishes variants known to cause ReNU syndrome from those in population controls. We redefine the CR at single-nucleotide resolution, resolve pathogenicity assignments for variants of uncertain significance, and show that function scores for variants within the CR correlate closely with phenotypic severity. Furthermore, we identify functionally critical variants in other regions of *RNU4-2* that underlie a recessive NDD marked by clinical features that are distinct from those of ReNU syndrome.

## RESULTS

### An optimised SGE assay reveals the functional spectrum of RNU4-2 variants

Performing SGE on regions of high sequence homology poses a challenge in that the protocol requires CRISPR/Cas9 editing of a single locus, specific amplification of the edited locus from millions of cells, and accurate variant calling from amplicon sequencing. Alignment of *RNU4-2* (RefSeq: NR_003137.3) to *RNU4-1* (RefSeq: NR_003925.1) reveals mismatches at only four of the 145 nucleotides. The sequence upstream of *RNU4-2*, however, is both unique and poorly conserved across species, such that guide RNAs (gRNAs) predicted to be highly specific^15^ can be designed in conjunction with PAM-disrupting edits to block Cas9 re-cutting (**Fig. 1A**).

**Figure 1.**
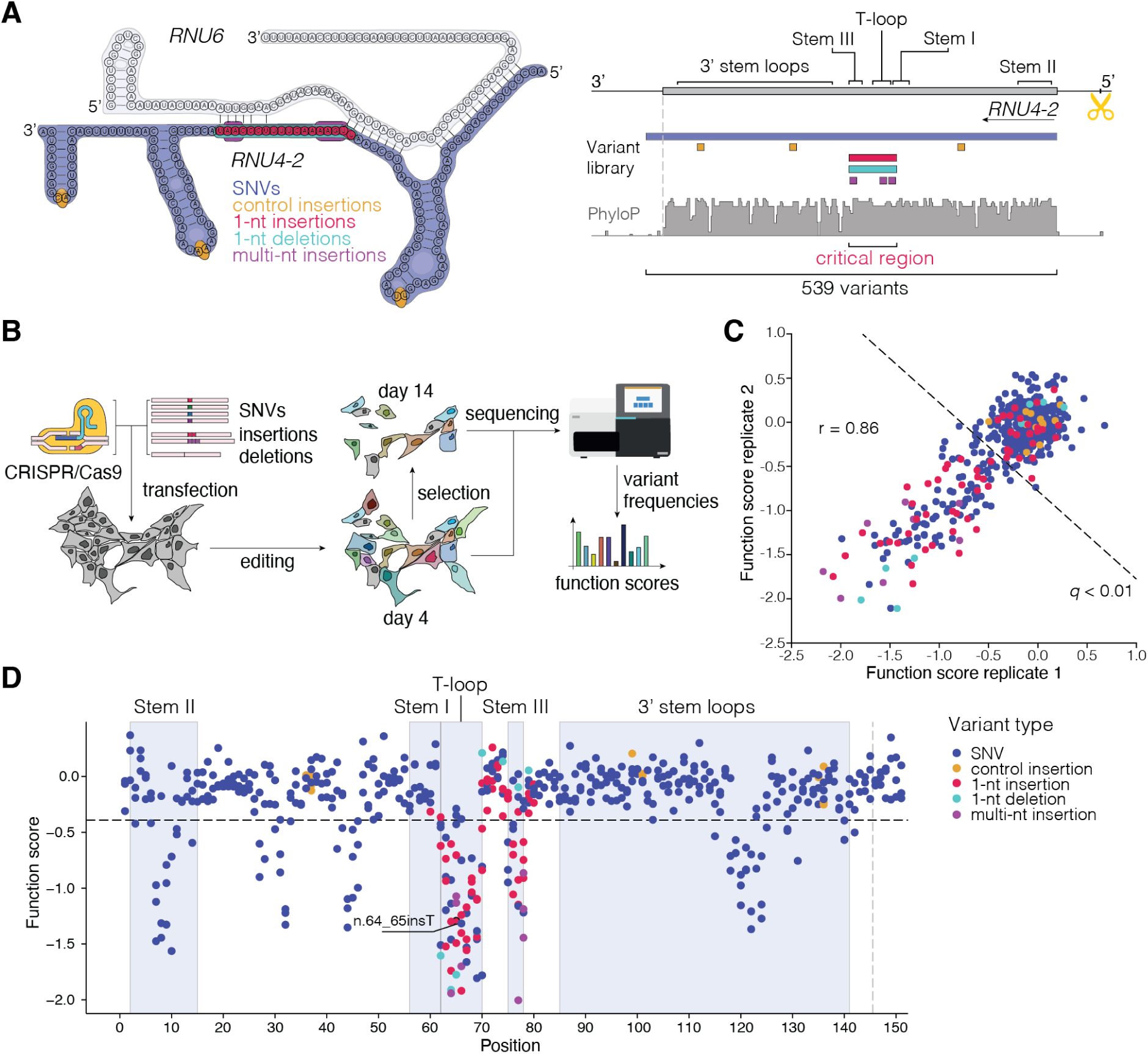
Saturation Genome Editing reveals the functional spectrum of *RNU4-2* variants. **A.** Schematic of SGE library design and CRISPR targeting strategy for *RNU4-2*. Positions of library variants including all possible SNVs (navy; across the 145-nt transcript and 6-nt 3’), control 1-nt insertions in loop regions (yellow), critical region (CR) 1-nt insertions (red) and deletions (teal) and multi-nt insertions (purple) are denoted on a schematic of *RNU4-2* and *RNU6* in complex (left) and by genomic location (right). A gRNA was designed to cleave upstream of *RNU4-2* (scissors), avoiding highly repetitive sequence and allowing for a PAM-blocking variant to be installed in a region of low conservation. (PhyloP 100 vertebrates basewise conservation track shown.) **B.** Schematic of SGE experiments in HAP1. Following editing, cells were harvested on days 4 and 14. Sequencing was performed to quantify variant frequencies at each timepoint and function scores were calculated. **C.** Function scores for 539 variants were correlated across biological replicates (Pearson’s r = 0.86). Variants scoring significantly lower than control insertions (*q* < 0.01) are indicated with the dashed line. **D.** Function scores are plotted by genomic position in relation to *RNU4-2* (RefSeq: NR_003137.3). The line at n.145 marks the end of the transcript, with 18 more distal SNVs also scored.

To perform SGE in HAP1, a haploid human line, we co-delivered Cas9 with a gRNA directing DNA cleavage 31-nt upstream of *RNU4-2* to install a library comprising 539 variants by homology-directed DNA repair (HDR). The library included all possible single base substitutions from the first transcribed nucleotide to six nucleotides beyond the most 3’ position of the *RNU4-2* transcript (GRCh38, chr12:120,291,753-120,291,903), as well as all 1-nt deletions and insertions in the CR, including all but one variant known to cause NDD (**Fig. 1A**). Uncertain whether pathogenic variants would display phenotypes in the HAP1-based assay, we included eight 2- to 5-nt insertions at positions in the CR previously associated with disease, reasoning these may have strong effects. As negative controls, we included twelve 1-nt insertions in stem loops outside the CR, which were not predicted to be deleterious (**Sup. Table 1**).

Adapting an optimized SGE protocol for HAP1 cells^13^ (**Fig. 1B**), we successfully scored all variants included in the library, observing an average of 64% editing by homology-directed repair (HDR) at day 4. Function scores, reflecting variants’ effects on growth (see **Methods**), were highly correlated across biological replicates (Pearson’s r = 0.86; **Fig. 1C**). As expected, given their location in the U4/U6 secondary structure, all twelve negative control variants scored near zero (mean = -0.03, s.d. = 0.13). We defined a neutral distribution from these negative controls to identify 138 significantly depleted variants (*q* < 0.01, i.e., function score < -0.39). The eight multi-nucleotide insertions in the CR included as positive controls all were depleted, with function scores ranging from -0.86 to -2.00. Mapping variants’ function scores to their linear transcript position reveals six distinct regions where variants score as depleted (**Fig. 1D**).

### SGE function scores resolve variant pathogenicity

We annotated all assayed variants within *RNU4-2* with whether or not they had been observed in individuals with ReNU syndrome^1,4^, observed in population cohorts (UK Biobank^16^ or All of Us), or observed in neither (unobserved; **Fig. 2A**). All 18 variants observed in ReNU syndrome were depleted in the assay (function score < -0.39), whereas 83.3% (290/348) of population variants scored as normal (function score ≥ -0.39; **Fig. 2B**). Accordingly, function scores effectively discriminate between ReNU syndrome variants and those identified in the population (**Fig. 2C**; area under the receiver operating characteristic (ROC) curve (AUC) = 0.95). Most variants that are unobserved in population cohorts score normally (60.0%; 93/155), however many are as, or even more, depleted than ReNU syndrome variants. Specifically, the five variants with the lowest function scores are all unobserved (**Sup. Table 1**).

**Figure 2.**
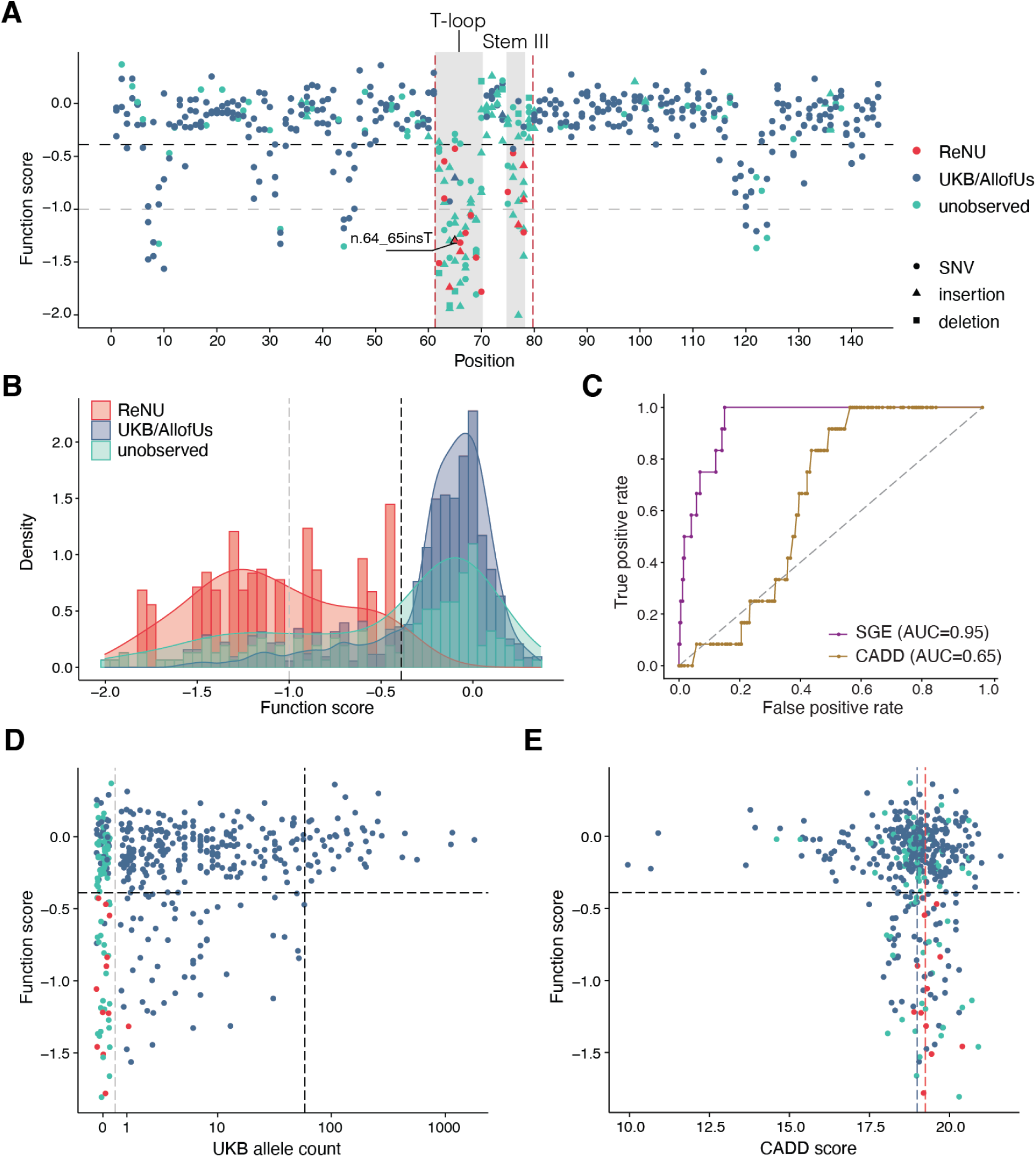
Function scores accurately discriminate variants underlying ReNU syndrome. **A.** Function scores for 521 variants within the *RNU4-2* transcript are plotted by position and coloured by their association with ReNU syndrome (red), presence in the UK Biobank or All of Us cohorts (blue), or no observation in either (teal). Depleted variants within the 18-nt CR (marked by vertical red dashed lines) are confined to two smaller regions (shaded grey) and include all ReNU syndrome variants scored (*n* = 18). These regions, n.62-70 and n.75-78, correspond to the T-loop and Stem III, respectively. The black dashed line (function score = -0.39) indicates significantly depleted variants and the gray dashed line (function score = -1.00) separates “moderate” from “strong” depletion. **B.** Stacked histogram and overlaid density plot of function scores by category comparing 18 ReNU syndrome variants to 348 variants observed in UK Biobank and/or All of Us and 155 unobserved variants. **C.** ROC curves show the performance of SGE function scores and CADD scores for classifying ReNU syndrome SNVs (*n* = 12) from SNVs observed at least once in population controls (*n* = 346). **D.** Function scores for SNVs are plotted by UK Biobank allele count (AC). Higher allele counts were correlated with higher function scores (Spearman’s ρ = 0.19, *P* = 5.3 x 10^-5^). Among 43 SNVs with allele count > 59 (black dashed line), no SNVs were depleted. The gray dashed line separates variants absent from UK Biobank (AC = 0) from those observed (AC > 0). **E.** Function scores for 435 SNVs are plotted by CADD score. The dashed line at y = -0.39 indicates significantly depleted SNVs, whereas the red line at x = 19.25 and the blue line at x = 18.99 indicate median CADD scores for ReNU syndrome SNVs and SNVs present in population cohorts, respectively.

We observed a significant correlation between SNV allele counts in UK Biobank and function scores, with rarer SNVs tending to be more depleted by SGE (Spearman’s ρ = 0.19, *P* = 5.3 x 10^-5^; **Fig. 2D**). Among 43 SNVs with an allele count of at least 60 in UK Biobank, none were depleted in the assay. Indeed, applying more stringent allele count thresholds to define control variants in population cohorts consistently improved the assay’s classification performance (**Sup. Fig. 1**). These findings indicate that depleted variants observed in population cohorts are unlikely to be the result of experimental noise and, instead, represent genuine variants impacting *RNU4-2* function segregating in the general population.

The discriminatory power of our SGE assay was substantially greater than that of the genome-wide *in silico* tool CADD^17^ (**Fig. 2C**; AUC = 0.65). Given the high conservation of the entire *RNU4-2* gene, most SNVs have very similar CADD scores (**Fig. 2E**). While CADD scores for ReNU syndrome SNVs are marginally higher on average than those for SNVs in population cohorts (NDD median = 19.2; UKB/All of Us mean = 19.0; one-sided Wilcoxon *P* = 0.040), a CADD score threshold that would capture all ReNU syndrome SNVs (≥ 18.89) would also annotate 56.4% (195/346) of SNVs observed in UK Biobank / All of Us, and 55.6% (190/342) of SNVs with normal SGE function scores, as likely deleterious. In contrast, our SGE function score threshold of -0.39 captures all ReNU syndrome SNVs and only 16.3% (56/343) of SNVs observed in population cohorts.

The assay clearly delineates the 18-nt CR of *RNU4-2* (**Fig. 2A**) within which variants cause ReNU syndrome, however, some variants in this region score normally. Using these data, we redefine the CR to two smaller regions of 9-nt (n.62-70, inclusive of insertions at n.61_62) and 4 nt (n.75-78), corresponding to the T-loop and Stem III, respectively (**Sup. Fig. 2**). Within these two regions, 84.3% (75/89) of tested variants (76.9% of SNVs), including all ReNU syndrome variants, have significant function scores, compared to 14.6% (63/432) across the remainder of *RNU4-2*.

Recent work by Nava *et al*.^4^ classified three variants outside of the CR and one deletion within the CR as variants of uncertain significance (VUS). Three of these variants were included in our assay (n.76del, n.92C>G, and n.111C>T) and all three had normal function scores (0.13, -0.08, and 0.09, respectively). Notably, all three variants are also observed in population controls. Further, a recent paper proposed a link between two 5’ stem loop variants, each identified in a single individual and inherited from an unaffected mother, and ReNU syndrome^5^. One of these variants is included in our assay (n.30A>T); its score of -0.25 is within the normal range, suggesting that this variant is unlikely to cause ReNU syndrome. Finally, of two variants recently associated with retinitis pigmentosa^18^, the one that is included in our assay (n.56T>C) also has a normal function score (-0.26).

### SGE depletion correlates with NDD severity

Nava *et al*. proposed a difference in phenotypic severity between ReNU syndrome variants mapping to the T-loop and Stem III structures of the U4/U6 duplex^4^. This difference is seen in our data, with Stem III variants having on average, higher function scores (T-loop mean = -1.22; Stem III mean = -0.86; one-sided Wilcoxon *P* = 0.033). However, we also observe considerable variation in function scores for NDD variants within each of the two regions. For example, two SNVs within the T-loop, n.63T>C and n.65A>G, have function scores above the mean observed for Stem III variants (-0.55 and -0.43, respectively). To investigate this, we repeated the phenotype clustering analysis of 143 individuals with ReNU syndrome from Nava *et al*.^4^ We classified the variants into two categories corresponding to ‘moderate’ (-1.0 < function score < -0.39) and ‘strong’ (function score < -1.0) levels of depletion in the assay (**Fig. 2A**; **Sup. Fig. 2**). All of the individuals with moderate category variants cluster together, including the four individuals with the n.63T>C (*n* = 1) and n.65A>G (*n* = 3) T-loop variants (**Fig. 3A**).

**Figure 3.**
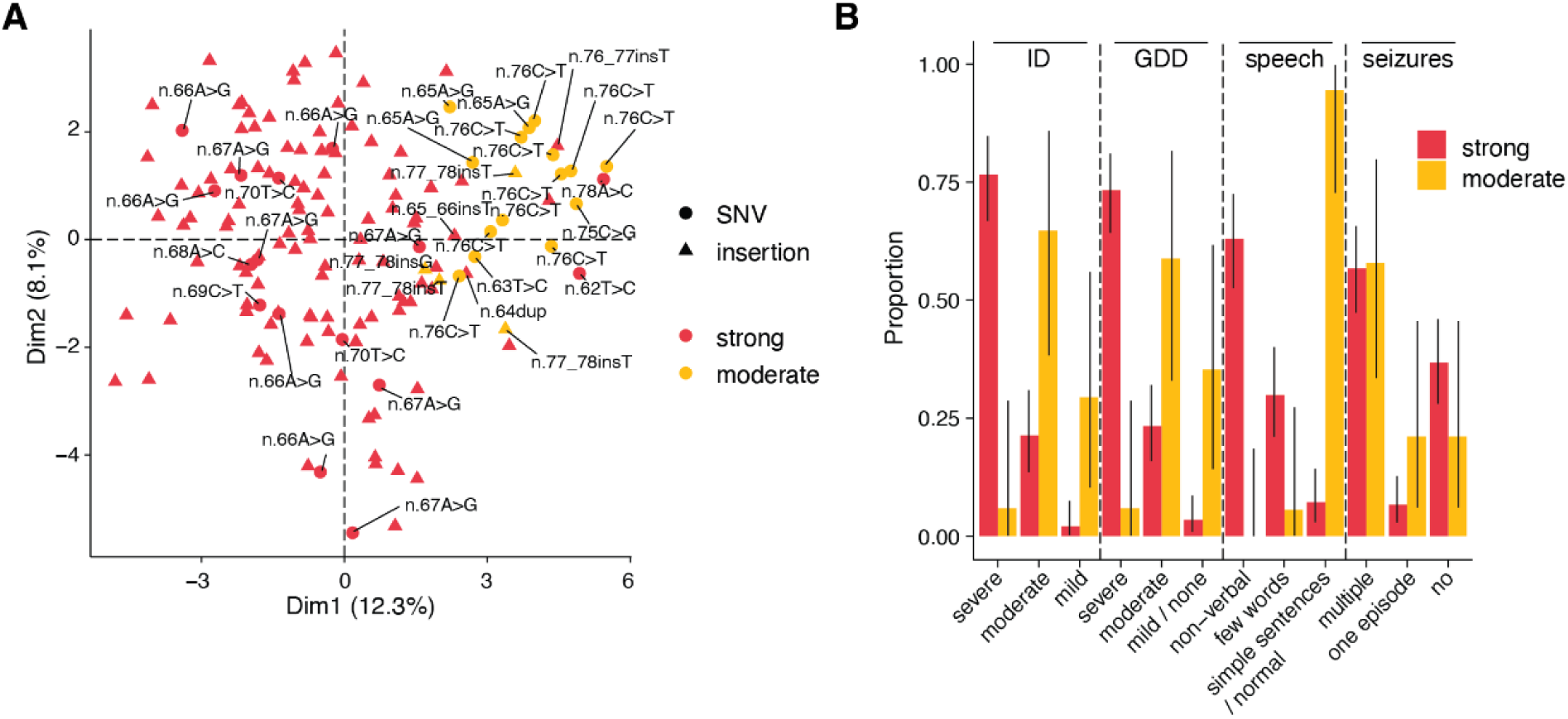
Function scores predict ReNU syndrome severity. **A.** The first two principal components from clustering of 143 ReNU syndrome cases by phenotype using the approach from Nava *et al*^4^. Variants are coloured by SGE function score class (strong depletion: function score < -1.0, moderate depletion: -1 < function score < -0.39). Unlabelled triangles indicate occurrences of n.64_65insT. **B.** The proportion of affected individuals with each phenotype is plotted, with cases grouped by SGE function score class. Error bars indicate 95% confidence intervals. ID: intellectual disability; GDD: global developmental delay.

To further determine if SGE function scores were able to discriminate between more severe and milder ReNU syndrome variants, we compared four specific phenotypes. Individuals with variants in the strong depletion group were significantly more likely to have severe developmental delay (73.3% vs 5.9%; OR = 42.7; 95%CI 6.1-1,841.8; two-sided Fisher’s *P* = 1.1×10^-7^), severe intellectual disability (76.6% vs 5.9%; OR=50.4; 95%CI 7.1-2,197.0; two-sided Fisher’s *P* = 3.6×10^-8^), and absent speech (92.8% vs 5.6%; OR=195.5; 95%CI 24.7-8,591.7; two-sided Fisher’s *P* = 6.6×10^-14^) than individuals with moderate depletion variants. There was no difference in the occurrence of seizures between variant groups (**Fig. 3B; Sup. Table 2**).

### A recessive NDD associated with variants in RNU4-2

Sixty-three variants outside of the T-loop and Stem III regions are depleted in the SGE assay (**Sup. Table 1**). Unlike the depleted variants in the ReNU CR, most of these other depleted variants (85.7%; 54/63) are observed in population control cohorts, albeit at low frequencies (**Fig. 2A**). To investigate whether these variants are associated with NDD-related traits, we compared individuals heterozygous for such variants (*n* = 50) and individuals with non-depleted SNVs (*n* = 12,134) in *RNU4-2* to individuals without any variants in *RNU4-2*, using the UK Biobank. We did not find any significant differences in fluid intelligence scores, childhood developmental disorder diagnoses, or age of leaving education (**Sup. Table 3**).

Because our SGE assay was performed in a haploid cell line, we reasoned that depleted variants outside of the critical region may instead be associated with recessive phenotypes. We searched global rare disease cohorts and identified sixteen individuals with biallelic depleted variants: nine (including three pairs of siblings) with homozygous variants and seven (including three pairs of siblings) who were each concordant for compound heterozygous depleted variants (**Sup. Table 4**). None of these variants were located in the ReNU CR, yet all sixteen individuals had NDD phenotypes. Across the rare disease cohorts, no individuals with phenotypes unrelated to NDD had biallelic depleted variants. Only a single individual across the UK Biobank and All of Us cohorts is homozygous for a SGE-depleted variant (n.31T>G, function score = -0.696). This individual has only primary level education (Highest Grade: One Through Four) and reports difficulties with ‘dressing or bathing’, ‘doing errands alone’, and ‘concentrating, remembering, or making decisions’, consistent with a possible intellectual disability. In the sixteen identified individuals we defined a novel neurodevelopmental disorder characterised by global developmental delay, intellectual disability, delayed or absent speech, hypotonia, spasticity, microcephaly, ophthalmological and visual impairments, and seizures, with variable involvement of genitalia, skin, hair, and limb anomalies. On MRI, individuals show distinctive white matter abnormalities and cerebellar atrophy that are not seen in ReNU syndrome. The clinical phenotypes of these individuals are discussed further in an accompanying manuscript.

Depleted variants outside of the ReNU CR broadly map to four regions of U4 that are known to mediate interactions between U4 and other components of the spliceosome: (1) the central portion of the Stem II interaction with U6 from nt 6 to 11^3^; (2) a region from nt 118 to 126 that interacts with a ring of Sm proteins that are important for U4 biogenesis and stability^19,20^; and (3) two regions mapping to either side of a ‘k-turn’ structure required for protein binding^21,22^ from nt 27 to 33 and 42 to 46 (**Fig. 4A**). Variants in the equivalent regions of *RNU4ATAC*, which encodes the minor spliceosome equivalent of U4, U4atac, cause rare recessive RNU4atac-opathies^23–25^. Of the eleven unique *RNU4-2* variants identified in the recessive NDD cases, four have exact equivalents in *RNU4ATAC* that are (likely) pathogenic in ClinVar (n.32G>A, n.45G>C, n.46G>A, and n.119A>G; **Sup. Table 5**). They include n.119A>G (function score = -0.686; *RNU4ATAC* equivalent: n.117A>G; ClinVar variation ID: 1525441), which was homozygous in two individuals and compound heterozygous in two brothers.

**Figure 4.**
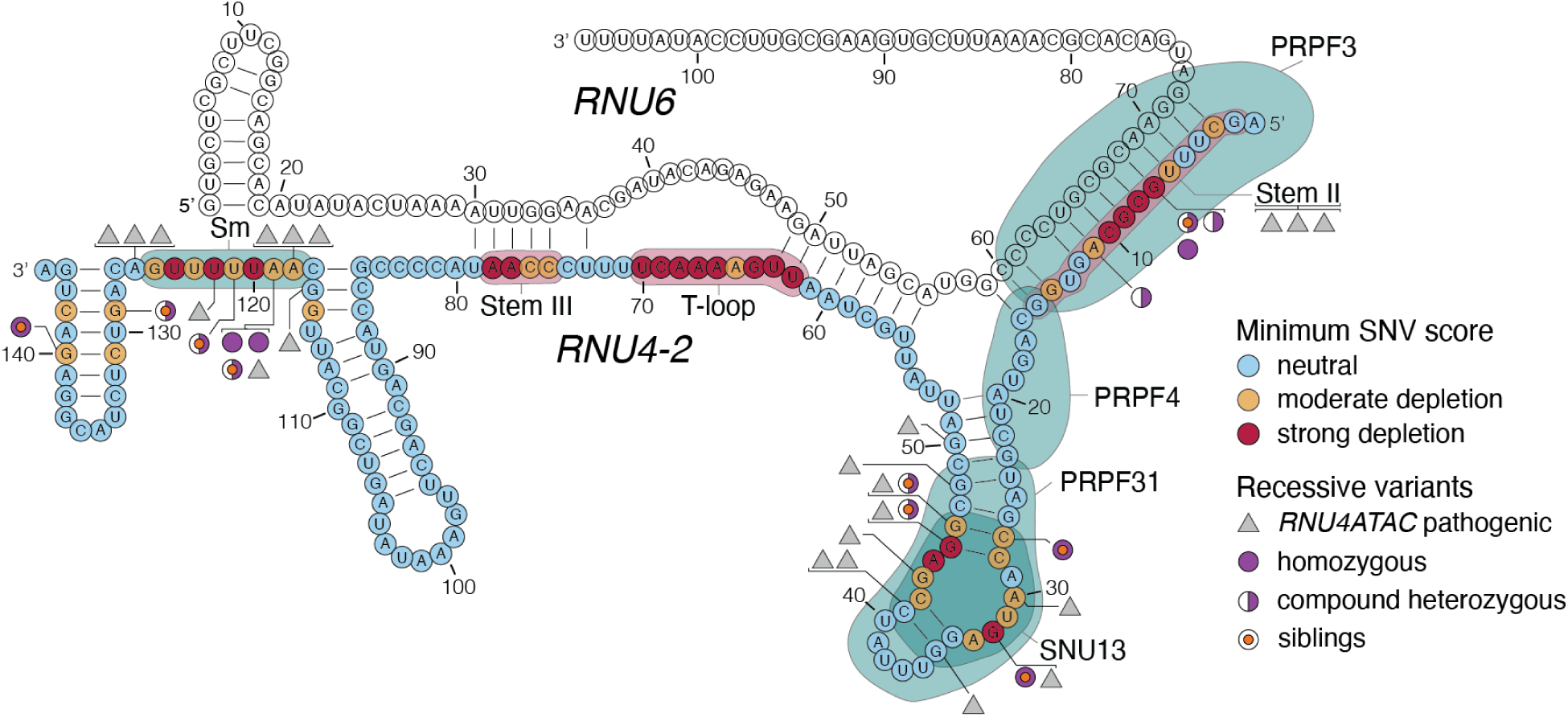
SGE-depleted variants outside the CR cause a recessive NDD. The lowest SGE function score class among SNVs at each position is indicated on the *U4*:*U6* secondary structure. Outside the CR, low SGE scores occur at positions of spliceosomal protein binding, indicated by teal shaded regions. Black triangles correspond to homologous positions of *RNU4ATAC* at which (likely) pathogenic variants have been linked to recessive disease (from ClinVar; **Sup. Table 5**). *RNU4-2* variants with low function scores observed in recessive NDD cases are indicated, with filled purple circles indicating variants observed as homozygous and half-filled circles indicating variants observed in the compound heterozygous state. An orange dot in the centre of a circle indicates that the variant is observed in two affected siblings. Six (likely) pathogenic *RNU4ATAC* variants could not be confidently assigned to an equivalent nucleotide in *RNU4-2*. Three of these (n.8C>A, n.13C>T, and n.16G>A) are shown together as mapping to Stem II. The other three (n.29T>G, n.30G>A, and n.111G>A) are not shown. U4/U6 structure depiction adapted from Quinodoz *et al.^18^*

## DISCUSSION

*RNU4-2* was the first non-coding RNA to be identified as having a substantial contribution to the prevalence of NDD, with ReNU syndrome predicted to impact around 100,000 individuals worldwide. Here, we developed an SGE assay to systematically assess the function of variants across *RNU4-2* and map genotype-phenotype relationships. We show that function scores accurately identify variants underlying ReNU syndrome and can distinguish these variants by disease severity. Further, we define the critical region at the centre of *RNU4-2* within which variants cause dominant ReNU syndrome, at nucleotide resolution. In two regions, of 9 nt and 4 nt, 84.3% of all tested variants are depleted. However, some variants in these regions, particularly in Stem III, have normal function scores and are therefore unlikely to be pathogenic. Consequently, these data have immediate utility in clinical interpretation of newly observed variants in individuals with NDD.

We identified four regions on the linear structure of *RNU4-2*, mapping to three regions of the U4/U6 duplex structure, outside of the ReNU CR where variants are also depleted. The same regions have previously been shown to be important in U4 mutational analyses in Yeast^20^. Given that the depleted variants are found in key regions of interaction between U4 and other important spliceosome factors, it is likely that they act by destabilising these interactions. This is supported for variants in the 5’ stem loop k-turn by previous work showing that the nucleotides that we identify as depleted are essential for SNU13/15.5k protein binding in vitro^22^. We hypothesised that these variants were depleted in SGE only because we used a haploid cell line, and may be damaging only when homozygous or compound heterozygous with another depleted variant. This led us to uncover a novel recessive NDD caused by homozygous and compound heterozygous variants in these regions that were depleted in SGE. This NDD is phenotypically distinct from ReNU syndrome. For example, MRI findings in individuals with ReNU syndrome most commonly include enlarged ventricles and corpus callosum abnormalities^4^, whereas individuals with biallelic *RNU4-2* variants commonly have progressive white matter changes and cerebellar atrophy (characterised thoroughly in a companion paper).

We expect that distinct mechanisms might underlie dominant and recessive *RNU4-2*-associated NDDs. We previously showed that individuals with ReNU syndrome have an increase in use of alternative non-canonical 5’ splice sites^1,4^, consistent with the role of the T-loop and Stem III regions in accurately positioning the U6 ACAGAGA sequence to receive the 5’ splice site. Recessive *RNU4-2* variants map to different locations within U4, outside of the T-loop and Stem III. As variants in the equivalent regions and nucleotides of *RNU4ATAC* that cause recessive RNU4atac-opathies have been shown to lead to intron retention^26,27^, a similar mechanism may underlie recessive *RNU4-2* NDD. This will need to be confirmed through RNA-sequencing of individuals with recessive disease^26,27^.

Intriguingly, most *RNU4-2* variants we identified in recessive NDD cases score as moderately depleted by SGE. As the major spliceosome is responsible for splicing the vast majority of introns, it is plausible that greater disruption of *RNU4-2* function is incompatible with life. As we have observed for CR variants associated with ReNU syndrome, the degree of functional impact caused by recessive NDD variants may correlate with disease severity. There may also be phenotypic differences between individuals with variants mapping to the three distinct regions we identified. Thorough phenotyping of large cohorts of cases will be necessary to establish how the degree of functional effect impacts phenotype. Additional SGE experiments incorporating larger insertions and deletions both inside and outside of the ReNU CR will add insights into the degree of tolerated disruption across different regions of *RNU4-2*. For example, Nava *et al*.^4^ identified a two-nt deletion (n.72_73del) in two individuals. This variant falls between Stem III and the T-loop but suggests that larger insertions and deletions in this region may also be disruptive to these structures.

Our HAP1-based SGE assay has several limitations. For one, we set a relatively conservative threshold to define significantly depleted variants (*q* < 0.01). While all variants associated with ReNU syndrome scored below this threshold, we cannot exclude the possibility that variants with more subtle effects may be clinically relevant. Secondly, the growth-based readout does not inform directly on underlying mechanisms of splice alteration (e.g. altered 5’ splice site usage, intron retention). This means that in the haploid context, both dominant and recessive effects are observed which cannot be separated by function score alone. Additionally, specific changes in splicing underlying certain clinical phenotypes may not occur in HAP1 due to differences between cell types. It is notable, for instance, that a variant recently associated with retinitis pigmentosa (n.56T>C) did not score significantly. Furthermore, most individuals with ReNU syndrome (70-75%) have the same single base insertion, n.64_65insT. Our data indicate that this variant is not unique in its functional severity, with many variants scoring similarly, or having even lower function scores. This result could argue against high recurrence being the result of a particularly damaging functional effect driving ascertainment; however, we cannot rule out the possibility that this variant leads to unique changes in splicing not reflected in SGE function scores. Future experiments using additional cell types will be valuable for delineating mechanisms of *RNU4-2* pleiotropy.

In summary, this work illustrates the power of a variant effect map for a locus recently implicated in disease to discover new genotype-phenotype associations and understand mechanisms underlying disease. SGE data for *RNU4-2* will be critical for accurately diagnosing patients with currently unexplained NDD and provide insights that are valuable for efforts to design effective therapies. Finally, the SGE strategy we used to overcome the high sequence homology of *RNU4-2* can be replicated to dissect other snRNAs recently linked to disease^4,28,29^.

## METHODS

### Single guide RNA design and cloning

The gRNA used for SGE was designed using Benchling’s CRISPR design tool to search the *RNU4-2* locus, including upstream and downstream regions of low sequence homology to *RNU4-1* and pseudogenes, identifying a candidate with high on-target and low off-target scores. The gRNA spacer sequence was ligated into the pX459 backbone as previously described^30^. Briefly, complementary primers containing the spacer were ordered from IDT (**Sup. Table 6**), phosphorylated, hybridised, and ligated into the pX459 linearized backbone followed by PlasmidSafe DNAse (Lucigen) digestion. Next, 2 µl of the ligation reaction were transformed in NEB Stable Competent *E. coli* cells using the high-efficiency transformation protocol and 75 µl of transformant was plated on ampicillin-resistant plates and cultured overnight at 30°C. Three colonies were then picked and grown overnight at 37°C in 7 ml Lysogeny Broth (LB) supplemented with carbenicillin (100 µg/ml). Plasmid DNA was extracted using the QIAprep Spin Miniprep kit (Qiagen) and verified using Plasmidsaurus whole-plasmid sequencing. The selected clone was then grown in 100 ml of LB broth at 37°C in a shaking incubator supplemented with carbenicillin. The cells were then pelleted and the plasmid was extracted using a ZymoPure Maxiprep kit (Zymo research), endotoxins were removed using EndoZero columns (Zymo research) and the product was quantified with the Qubit dsDNA BR assay kit (Invitrogen).

### HDR library cloning

An oligonucleotide library comprising *RNU4-2* variants was manufactured by Twist Bioscience and subsequently cloned into a vector containing homology arms for *RNU4-2* to make the HDR library for SGE.

To generate the vector with homology arms, a nested PCR was performed on genomic DNA (gDNA) extracted from HAP1-LIG4-KO cells^10^ using primers designed to generate homology arms of 700 to 800 bp flanking *RNU4*-2 (**Sup. Table 6**). The PCR was performed using the Kapa HiFi HotStart ReadyMix (Roche). The product was purified using Ampure XP (Beckman Coulter) magnetic beads at 1.2x volume and eluted in 12 µl of nuclease-free water. The amplicon containing *RNU4-2* homology arms was then inserted in the linearized pUC19 backbone using In-Fusion HD cloning (Takara) and 2 µl of cloning reaction was transformed into NEB Stable cells following the manufacturer’s 5-minute transformation protocol. Cells were plated on agar plates containing ampicillin and incubated at 30°C overnight. The pUC19 plasmid containing *RNU4-2* homology arms (pUC19-RNU4-2-HA) was purified and sequence-verified from a successfully transformed clone. pUC19-RNU4-2-HA was then diluted to 8.7 pg in a 50 µl PCR reaction and amplified with Kapa HiFi to obtain a linearised product with 17-18 bp complementarity to the *RNU4-2* oligo library. A PAM-blocking mutation was introduced 27 nt upstream of the coding sequence (chr12:120,291,930-C-G) via primer overhang extension during PCR. The reaction was treated with 1 µl of DpnI (NEB) for 30 minutes at 37°C, gel extracted and quantified. Then, the *RNU4-2* oligo library was amplified using Kapa HiFi and purified using AmpureXP (1.2x). The amplified library and linearised pUC19-RNU4-2-HA plasmid were then assembled using the In-Fusion HD cloning kit, and the product was transformed into NEB stable cells using the high-efficiency transformation protocol. To quantify efficiency, 1% of cells in the transformation reaction were plated and the remainder were cultured in 100 ml LB with carbenicillin overnight at 37°C. Cells were then pelleted by centrifugation and the final *RNU4-2* HDR library was extracted using the ZymoPure Maxiprep kit (Zymo Research) with endotoxin removal. The isolated HDR library was quantified with a Qubit dsDNA BR assay kit and sequence-verified via Plasmidsaurus.

### HAP1 cell culture

HAP1*-*LIG4-KO cells (referred to throughout simply as ‘HAP1’) display increased rates of editing by HDR due to a frameshifting mutation in *LIG4^10^*. Frozen HAP1 cells were thawed at 37°C in a water bath, then supplemented with 10 ml of pre-warmed Iscove’s Modified Dulbecco’s Medium (IMDM) containing L-glutamine, 25 nM HEPES (Gibco), 10% FBS (Gibco), 1% Penicillin–Streptomycin (Gibco) and 2.5 μM 10-deacetyl-baccatin-III (DAB, Stratech), herein referenced to as IMDMc. Cells were centrifuged at 300 rcf for 3 minutes. The supernatant was then aspirated, and the cells were resuspended in fresh media, plated on a 10-cm dish, and cultured at 37°C with 5% CO_2_. The next day, the IMDMc media was replaced, and cells were cultured routinely from that point forward.

The HAP1 sub-culture routine included a 1:5 split every 48 hours or 1:10 split every 72 hours, to prevent cells from exceeding 80% confluency. To split cells, the media was aspirated and the dish washed with 10 ml of room temperature DPBS (Gibco). Following DPBS aspiration, the cells were treated with 1 ml of 0.25% trypsin–EDTA (Gibco) and incubated for 3 minutes at 37°C. 14 ml of pre-warmed IMDMc was then added and cells were collected and centrifuged at 300 rcf for 5 minutes. Cells were then re-suspended in 10 ml IMDMc, counted and seeded on a 10-cm dish.

### Transfection and selection

The day before transfection, 12 million cells were seeded on a 10-cm dish for each replicate and 2 million cells were seeded on a 6-well plate for the negative control sample. On the day of transfection (day 0), a transfection mix containing 10 µg of HDR library, 30 µg of the pX459 gRNA plasmid and 24 µl of Xfect polymer (Takara) in a final volume of 800 µl was prepared according to the manufacturer’s instructions for each replicates. For the negative control sample, a pX459 plasmid with a gRNA targeting *HPRT1^13^* instead of *RNU4-2* was used to prevent successful editing, and the transfection volume mix was scaled down 8-fold. Following transfection, cells were incubated for 24 hours at 37°C and supplemented with pre-warmed IMDMc with 1 µg/ml puromycin (Cayman Chemical). On day 4, half of the cells for each replicate were harvested for gDNA extraction and stored as a pellet at -70°C; the rest were kept in culture in 15-cm dishes supplemented with 15 ml of IMDMc. The negative control sample was harvested when reaching 70% confluency at day 6. A second sample of 10 million cells per replicate was collected at day 14 and stored at -70°C.

### Sequencing library preparation

gDNA was extracted from cells using QIAshredder (Qiagen) columns followed by the Allprep DNA/RNA kit (Qiagen) according to the manufacturer’s instructions. Concentrations were determined using the Qubit dsDNA BR assay kit. The *RNU4-2* locus was subsequently amplified using nested PCR to avoid amplification of plasmid DNA, followed by an indexing PCR, in total using three primer sets (**Sup. Table 6**). For the first reaction, the total gDNA template from each condition was partitioned into separate reactions, each containing 1.25 µg of DNA in a 100 µl reaction volume, using NEBNext Ultra II Q5 master mix (NEB) supplemented with MgCl_2_ (Ambion) to a final concentration 4 mM. The amplification reaction was monitored by qPCR using SYBR green (Invitrogen) and stopped prior to completion. The reactions for each sample were pooled and mixed before 50 µl of each product was purified using AmpureXP (1.2x) and eluted in 15 µl of nuclease-free water. 1 µl of purified product was loaded into the second qPCR reaction (50 µl final volume) and amplified using NEBNext Ultra II Q5. The reaction was again monitored using SYBR green and stopped prior to completion. The AmpureXP purification was then repeated, and a final qPCR (NEBNext Ultra II Q5) to incorporate sample indexes and sequencing adapters was performed using 1 µl of purified product as template in a 50 µl reaction for 8 cycles. Final products were purified and quantified with the Qubit dsDNA HS kit. The samples were then pooled for sequencing, aiming for 5 million reads per experimental replicate timepoint, 2 million reads for the negative control sample, and 1 million reads for the HDR library. The pool was purified using AmpureXP (1x), quantified, and loaded on a Novaseq X sequencer (Illumina).

### Variant frequency quantification

The fastq files were de-multiplexed using the *bcl2fastq* script and the variants were quantified as previously described^13^. Briefly, paired-end reads were adapter trimmed and merged, and reads containing N bases were discarded. HDR editing rates were computed from fastq files directly as the fraction of reads containing the exact PAM-blocking mutation. Fastq files were then aligned to a reference *RNU4-2* sequence and the frequency of each variant included in the library was determined.

### Function score calculation

All variants were observed in the library and day 4 at a frequency higher than 10^-4^, and were therefore included in downstream analyses. Function scores for library variants were first calculated per replicate, computed as the log_2_-ratio of day 14 to day 4 variant frequencies, normalised such that the median function score of negative control insertions equalled 0. Final function scores were then calculated for each variant by averaging function scores across replicates, again normalising to the median of negative control insertions. For each variant, p-values were determined using the *norm.cdf* function in Python, defining a normal distribution from the mean and standard deviation of function scores for negative control insertions. The p-values were corrected for multiple hypothesis testing using the *multipletests* function in Python (Benjamini-Hochberg procedure) to derive q-values. Significantly depleted variants were defined as those with q-values < 0.01, corresponding to a function score < -0.39. We further classified depleted variants into two categories using an arbitrary function score threshold of -1.0 to include sufficient variants and individuals per category to assess for phenotypic differences.

### Variant annotation

Variants were annotated as ReNU syndrome variants if they were reported in Chen *et al*.^1^ or classified as pathogenic or likely pathogenic in Nava *et al*.^4^. Variants were annotated with whether or not they were observed in the 490,640 genome sequenced individuals from the UK Biobank^16^ (DRAGEN pipeline) or in 414,840 individuals from All of Us V8. CADD v1.7^7^ annotations were obtained by uploading a synthetic VCF of all possible SNVs to the online annotation tool (https://cadd.gs.washington.edu/score). As we pre-selected which insertions and deletions to include in the SGE assay (due to assay size limitations), we restricted analyses involving CADD to SNVs within the *RNU4-2* transcript.

Receiver operating characteristic (ROC) area under the curve (AUC) values were calculated by assigning a “1” label to ReNU syndrome SNVs and a “0” label for SNVs observed in UK Biobank or All of Us. The labels and corresponding function scores were used to compute false positive and true positive rates (using Python’s *roc_curve* function), as well as ROC-AUC values (using the *roc_auc_score* function). This analysis was also restricted to SNVs only.

### Phenotype severity and clustering

Categorical data for 44 clinical features from 143 patients with pathogenic and likely pathogenic *RNU4-2* variants^4^ were transformed into a 0-1 scale, with 0 indicating a more favourable phenotype and 1 a more severe presentation. PCA was generated after imputing missing data with 0 and performing variable scaling. Two-sided Fisher’s tests with Bonferroni adjustment to account for four tests were used to compare clinical features between SGE function score variant categories (strong vs moderate) in **Sup. Table 2**.

### Association testing in UK Biobank

We extracted phenotypes associated with educational attainment from UK Biobank following an approach published previously^31^. Fluid intelligence scores (field ID:20016) were retrieved for all participants. Where multiple scores were recorded, the median value was taken. Age left education was calculated as the maximum value in age completed full time education (field ID: 845). Diagnosis with childhood developmental disorder (child DD) was defined using the ICD codes for intellectual disability (ICD-10: F70-F73, F78, F79; ICD-9: 317, 318, 319), epilepsy (ICD-10: G40), global developmental disorders (ICD-10: F80-F84, F88-F95, R62, R48, Z55; ICD-9: 299, 312, 313, 314, 315), and congenital malformations (ICD-10: Q0-Q99, ICD-9: 740-759).

We identified UK Biobank participants with: (1) depleted variants in the 18 bp *RNU4-2* CR (*n* = 6), (2) depleted variants outside of the CR (*n* = 50), and (3) participants with non-depleted SNVs outside of the CR (*n* = 12,132). We performed multiple linear regression on fluid intelligence scores and age left education, and multiple logistic regression on child DD for variant groups (2) and (3) defined above, compared to all individuals without any variants in any of the three groups. Age at recruitment (field ID:21022), age^2 (age at recruitment * age at recruitment), sex (field ID: 31), and first ten genetic principal components (field ID:22009) were included as covariates. *P*-values were FDR corrected using the Benjamini-Hochberg method.

### Investigating RNU4ATAC variants in ClinVar

Variants in *RNU4ATAC* with classifications of pathogenic, likely pathogenic, pathogenic/likely pathogenic, benign, likely benign or benign/likely benign were downloaded from the ClinVar^32^ website on 4th March 2025. Two regions of *RNU4-2* and *RNU4ATAC* with identical structures were defined, mapping to the k-turn (*RNU4-2* nt 26-52; *RNU4ATAC* nt 31-57) and the Sm protein binding site (*RNU4-2* nt 115-126; *RNU4ATAC* nt 113-124). Variants at the same nucleotide in the structure and where the reference bases in *RNU4-2* and *RNU4ATAC* are identical, were marked as ‘equivalent’.

### Identifying biallelic variants in rare disease and population cohorts

We searched rare disease cohorts for individuals with biallelic variants in *RNU4-2*. These cohorts included the Genomics England 100,000 Genomes Project and NHS Genomic Medicine Service datasets accessed through the UK National Genomic Research Library^33^, the SeqOIA and Auragen clinical cohorts in France (PFMG 2025), the Undiagnosed Disease Network, the Broad Institute Center for Mendelian Genomics (CMG) and GREGoR (Genomics Research to Elucidate the Genetics of Rare Diseases)^34^ Consortium cohorts. We only included individuals with homozygous variants with function scores < -0.39, or compound heterozygous variants where both had function scores < -0.39.

Informed consent was obtained for all patients included in this study from their parent(s) or legal guardian, with the study approved by the local regulatory authority. The 100,000 Genomes Project Protocol has ethical approval from the HRA Committee East of England Cambridge South (REC Ref 14/EE/1112).

We received an exception to the Data and Statistics Dissemination Policy from the All of Us Resource Access Board to report questionnaire response data for the single individual with a homozygous depleted variant as well as variant counts < 20 for all variants in *RNU4-2*.

## DATA AVAILABILITY

SGE data including all *RNU4-2* function scores are available in **Sup. Table 1**. Fastq files from SGE experiments are available on the European Nucleotide Archive (accession: PRJEB87505).

## CODE AVAILABILITY

Custom scripts used to analyse SGE experiments and generate figures are available on GitHub: https://github.com/FrancisCrickInstitute/RNU4-2_Saturation_Genome_Editing.

## AUTHOR CONTRIBUTIONS

J.D.J. and A.A. performed experiments. J.D.J., H.C.K., R.D., E.L., Y.C., A.B., and J.L., analysed data and contributed to the figures and tables in the paper. C.S., R.R., H.R.M., D.G.M., C.D., N.W., and G.M.F. collected data, provided funding and supervised the work. All other authors provided clinical and/or genomic data and are listed alphabetically. J.D.J, N.W., and G.M.F. wrote the paper with input from all the authors.

## ACKNOWLEDGEMENTS AND FUNDING

We thank the Crick’s Genomics Scientific Technology Platform (STP) for performing sequencing and the Cell Services STP for assisting in maintaining cell lines. We also thank Peter O’Donovan, Mitra Sato and Eve Miller from the Genomics England Airlock team.

N.W. is supported by a Sir Henry Dale Fellowship jointly funded by the Wellcome Trust and the Royal Society (grant no. 220134/Z/20/Z), a Lister Institute research prize, and grant funding from Novo Nordisk. Y.C. is supported by a studentship from Novo Nordisk. The Francis Crick Institute receives its core funding (G.M.F.) from Cancer Research UK (CC2190), the UK Medical Research Council (CC2190), and the Wellcome Trust (CC2190). A.B. is supported by a Wellcome PhD Training Fellowship for Clinicians and the 4Ward North PhD Programme for Health Professionals (223521/Z/21/Z). C.D. is supported by research grants from the Deutsche Forschungsgemeinschaft (DFG) grant (Project numbers 455314768, 458099954, and 505514143). C.N. has received support from the Health philantropic program of Mutuelles AXA dedicated to supporting innovative research projects in France (RNU-SPLICE project). Patients 4, 5, 6, 13, 14, 15, and 16 included in this study were diagnosed through Plan France Médecine Génomique 2025 (PFMG2025). Patients 11 and 12 were sequenced at the Baylor College of Medicine Human Genome Sequencing Center through the GREGoR Consortium with support from US National Human Genome Research Institute grants U01HG011758 and U54HG003273. Analysis of individuals 9 and 10 was supported by NHGRI grant R01HG009141. D.G.C. was supported by the Child Neurologist Career Development Program CNCDP-K12 (US National Institute of Neurological Disorders and Strokes grant NS098482). C.A-T. is supported in part by the National Human Genome Research Institute (NHGRI) grant U01HG011755 (GREGoR consortium). O.M. is supported by the Hazem Ben-Gacem Tunisia Medical Fellowship Fund. Research reported in this publication was supported by the National Institute Of Neurological Disorders And Stroke of the National Institutes of Health under Award Numbers U01HG010218 and U01HG010233. The content is solely the responsibility of the authors and does not necessarily represent the official views of the National Institutes of Health. This research was made possible through access to data in the National Genomic Research Library, which is managed by Genomics England Limited (a wholly owned company of the Department of Health and Social Care). The National Genomic Research Library holds data provided by patients and collected by the NHS as part of their care and data collected as part of their participation in research. The National Genomic Research Library is funded by the National Institute for Health Research and NHS England. The Wellcome Trust, Cancer Research UK and the Medical Research Council have also funded research infrastructure. This study was registered with Genomics England under Research Registry Projects 354. This research has been conducted using the UK Biobank Resource under application number 81050. We gratefully acknowledge All of Us and UK Biobank participants for their contributions. We also thank the National Institutes of Health’s All of Us Research Program for making available the participant and variant data examined in this study.

For the purpose of Open Access, the authors have applied a CC BY public copyright license to any Author Accepted Manuscript version arising from this submission.

## SUPPLEMENTARY FIGURES

**Supplementary Figure 1:**
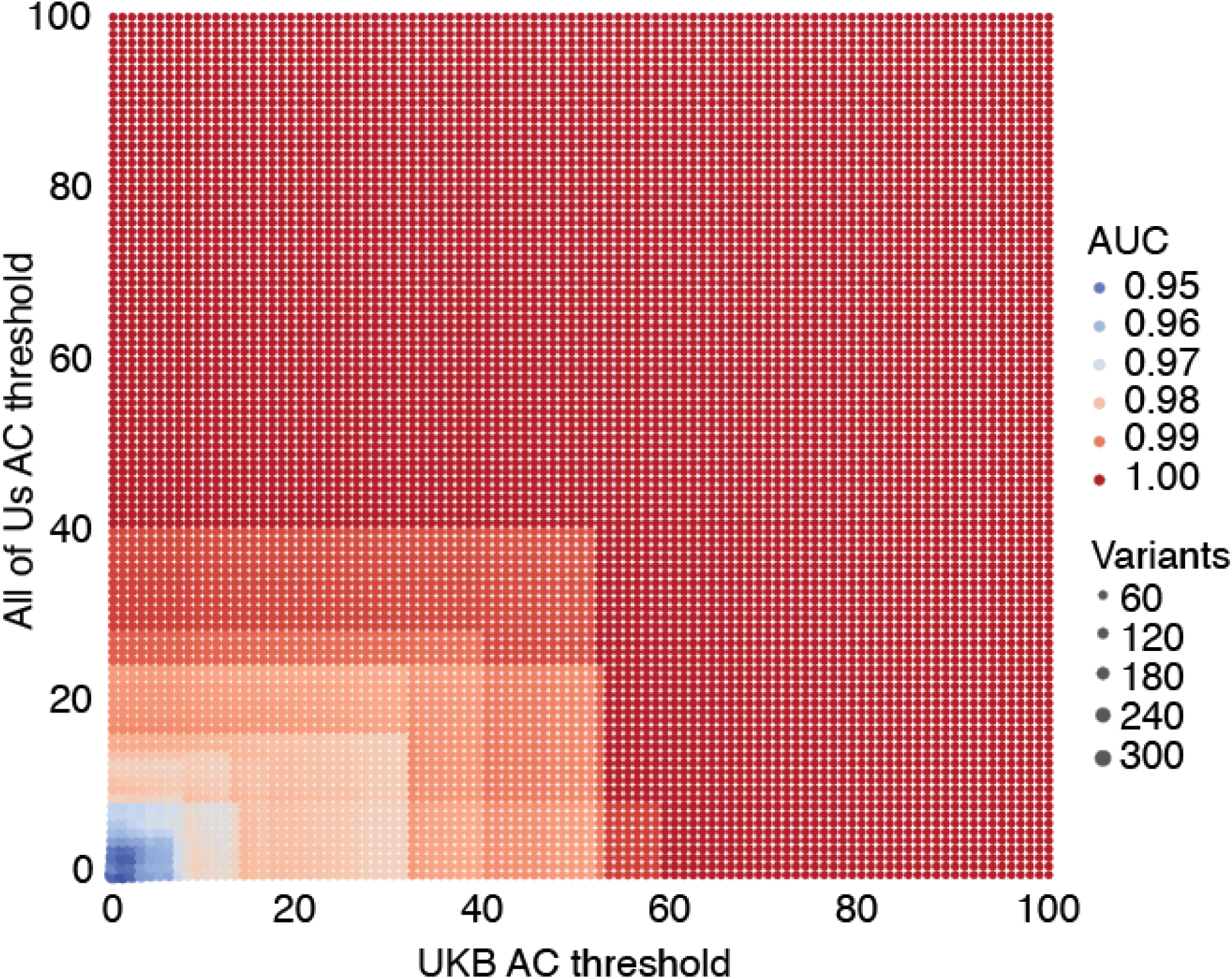
ReNU syndrome variants are discriminated with high precision from variants seen frequently in population controls. ROC-AUC measurements for distinguishing 12 ReNU syndrome SNVs from population control SNVs by SGE score are displayed as a heatmap. Each AUC was determined using only variants in UK Biobank and All of Us with allele counts above the thresholds indicated on the axes. Dot size indicates the number of population cohort variants retained in each AUC calculation.

**Supplementary Figure 2:**
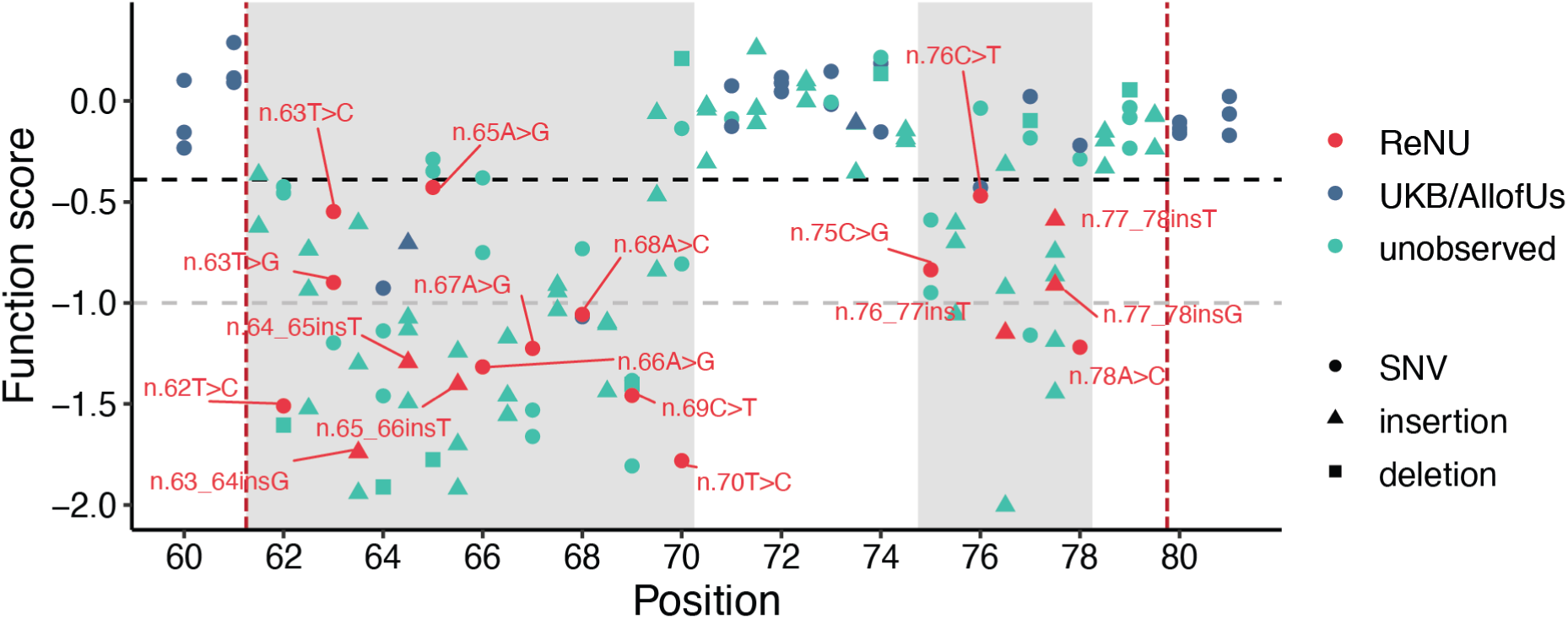
Function scores for variants within the *RNU4-2* critical region. Function scores are plotted by position and coloured by their association with ReNU syndrome (red), presence in the UK Biobank or All of Us cohorts (blue), or no observation in either (teal). Variants score lowly in two regions within the CR (shaded), n.62-70 and n.75-78, which correspond to the T-loop and Stem III, respectively. The black dashed line (function score = -0.39) indicates significantly depleted variants and the gray dashed line (function score = -1.00) separates “moderate” from “strong” depletion.

## SUPPLEMENTARY TABLES

**Supplementary Table 1: Details and function scores for *RNU4-2* variants assayed with SGE.** (provided as separate file)

**Supplementary Table 2:** Comparison of clinical features by function score categories. Two-sided Fisher’s tests were used to compare intellectual disability (severe vs mild/moderate), developmental delay (severe vs moderate/mild/none), speech ability (non-verbal/few words vs simple sentences/normal speech), and epilepsy/seizures (yes/one episode vs no) between strong vs moderate depleted variants in the SGE assay. P-values are Bonferroni adjusted for four tests.

| Phenotype | Category | Strong | Moderate | OR (95%CI) | P-value |
| --- | --- | --- | --- | --- | --- |
| Intellectual disability | Severe | 72/94 (0.766) | 1/17 (0.059) | 50.4 (7.1-2197.0) | 1.45x10<sup>-7</sup> |
|  | Moderate / mild | 22/94 (0.234) | 16/17 (0.941) |  |  |
| Global developmental delay | Severe | 85/116 (0.733) | 1/17 (0.059) | 42.7 (6.1-1841.8) | 4.28x10<sup>-7</sup> |
|  | Moderate / mild / none | 31/116 (0.267) | 16/17 (0.941) |  |  |
| Speech | Non verbal / few words | 90/97 (0.928) | 1/18 (0.056) | 195.5 (24.7-8591.7) | 2.66x10<sup>-13</sup> |
|  | Simple sentences / normal speech | 7/97 (0.072) | 17/18 (0.944) |  |  |
| Seizures | Yes, more than one episode | 68/120 (0.567) | 11/19 (0.579) | 0.46 (0.11-1.58) | 0.829 |
|  | One episode | 8/120 (0.067) | 4/19 (0.211) |  |  |
|  | No | 44/120 (0.367) | 4/19 (0.211) |  |  |

**Supplementary Table 3:** Results from association testing with intelligence-related metrics in the UK Biobank. Individuals with depleted variants outside of the ReNU syndrome critical region (non-CR; max *n* = 50), and individuals with SNVs with normal SGE function scores (≥ -0.39; max *n* = 12,132) were compared to individuals without variants in *RNU4-2* (fluid_intelligence *n* = 207,458; age left education *n* = 311,233, childhood developmental disorder (DD) = 453,754). The n shown in the table represents the number of individuals in each variant group without missing data that were included in each test.

| Phenotype | Variant group | n | Regression type | Coefficient (95% CI) | P-value |
| --- | --- | --- | --- | --- | --- |
| Fluid intelligence | depleted non-CR | 20 | linear | 0.366 (-0.523-1.256) | 0.420 |
|  | not depleted SNVs | 5,341 |  | -0.024 (-0.079-0.031) | 0.389 |
| Age left education | depleted non-CR | 34 | linear | -0.120 (-0.891-0.652) | 0.761 |
|  | not depleted SNVs | 8,003 |  | 0.012 (-0.040-0.062) | 0.678 |
| Child DD | depleted non-CR | 50 | logistic | -0.693 (-2.673-1.287) | 0.493 |
|  | not depleted SNVs | 12,134 |  | -0.041 (-0.136-0.054) | 0.393 |

**Supplementary Table 4:**
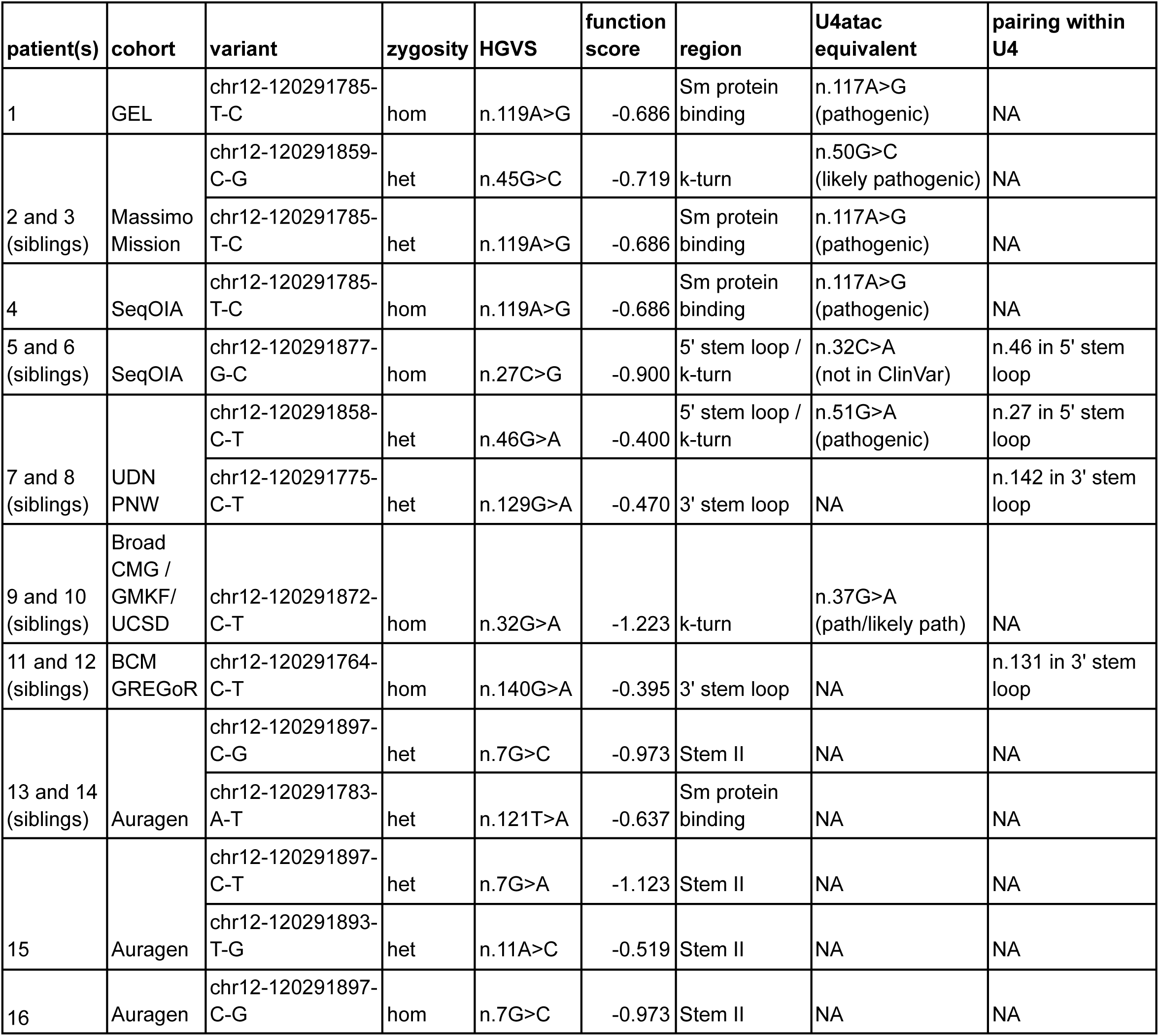
Homozygous and compound heterozygous variants in individuals with undiagnosed neurodevelopmental disorders. Equivalent variants in *RNU4ATAC* and their ClinVar classification are included for variants at the equivalent nucleotide and with the same reference (see **methods**). Where a variant is part of a stem region of pairing within the U4 structure, the base it pairs with is noted.

| patient(s) | cohort | variant | zygosity | HGVS | function score | region | U4atac equivalent | pairing within U4 |
| --- | --- | --- | --- | --- | --- | --- | --- | --- |
| 1 | GEL | chr12-120291785-T-C | hom | n.119A>G | -0.686 | Sm protein binding | n.117A>G (pathogenic) | NA |
| 2 and 3 (siblings) | Massimo Mission | chr12-120291859-C-G | het | n.45G>C | -0.719 | k-turn | n.50G>C (likely pathogenic) | NA |
|  |  | chr12-120291785-T-C | het | n.119A>G | -0.686 | Sm protein binding | n.117A>G (pathogenic) | NA |
| 4 | SeqOIA | chr12-120291785-T-C | hom | n.119A>G | -0.686 | Sm protein binding | n.117A>G (pathogenic) | NA |
| 5 and 6 (siblings) | SeqOIA | chr12-120291877-G-C | hom | n.27C>G | -0.900 | 5' stem loop / k-turn | n.32C>A (not in ClinVar) | n.46 in 5' stem loop |
| 7 and 8 (siblings) | UDN PNW | chr12-120291858-C-T | het | n.46G>A | -0.400 | 5' stem loop / k-turn | n.51G>A (pathogenic) | n.27 in 5' stem loop |
|  |  | chr12-120291775-C-T | het | n.129G>A | -0.470 | 3' stem loop | NA | n.142 in 3' stem loop |
| 9 and 10 (siblings) | Broad CMG / GMKF / UCSD | chr12-120291872-C-T | hom | n.32G>A | -1.223 | k-turn | n.37G>A (path/likely path) | NA |
| 11 and 12 (siblings) | BCM GREGoR | chr12-120291764-C-T | hom | n.140G>A | -0.395 | 3' stem loop | NA | n.131 in 3' stem loop |
| 13 and 14 (siblings) | Auragen | chr12-120291897-C-G | het | n.7G>C | -0.973 | Stem II | NA | NA |
|  |  | chr12-120291783-A-T | het | n.121T>A | -0.637 | Sm protein binding | NA | NA |
| 15 | Auragen | chr12-120291897-C-T | het | n.7G>A | -1.123 | Stem II | NA | NA |
|  |  | chr12-120291893-T-G | het | n.11A>C | -0.519 | Stem II | NA | NA |
| 16 | Auragen | chr12-120291897-C-G | hom | n.7G>C | -0.973 | Stem II | NA | NA |

**Supplementary Table 5:** List of variants in *RNU4ATAC* in ClinVar. The equivalent residue of RNU4-2 was determined for variants in the 5’ stem loop / k-turn and Sm protein binding site (see **methods**). Function scores are included where an equivalent nucleotide could be determined and the reference base at that position is the same across *RNU4-2* and *RNU4ATAC*.

| U4atac HGVS | Chr | Pos | ClinVar variation ID | ClinVar classification | U4 nt | matching ref? | function score | category | region |
| --- | --- | --- | --- | --- | --- | --- | --- | --- | --- |
| n.8C>A | 2 | 121530887 | 692040 | Likely pathogenic |  |  |  |  | Stem II |
| n.13C>T | 2 | 121530892 | 218083 | Pathogenic |  |  |  |  | Stem II |
| n.16G>A | 2 | 121530895 | 218082 | Pathogenic/Likely pathogenic |  |  |  |  | Stem II |
| n.29T>G | 2 | 121530908 | 977869 | Likely pathogenic |  |  |  |  | 5' stem loop |
| n.30G>A | 2 | 121530909 | 30180 | Likely pathogenic |  |  |  |  | 5' stem loop |
| n.35A>C | 2 | 121530914 | 2674599 | Likely pathogenic | 30 | Y | -0.4725 | UKB/AllofUs | 5' stem loop / k-turn |
| n.37G>A | 2 | 121530916 | 218084 | Pathogenic/Likely pathogenic | 32 | Y | -1.2228 | UKB/AllofUs | 5' stem loop / k-turn |
| n.40C>T | 2 | 121530919 | 599282 | Pathogenic | 35 | N |  |  | 5' stem loop / k-turn |
| n.46G>T | 2 | 121530925 | 977856 | Likely pathogenic | 41 | N |  |  | 5' stem loop / k-turn |
| n.46G>A | 2 | 121530925 | 636959 | Pathogenic | 41 | N |  |  | 5' stem loop / k-turn |
| n.48G>A | 2 | 121530927 | 218085 | Pathogenic/Likely pathogenic | 43 | Y | -0.1675 | UKB/AllofUs | 5' stem loop / k-turn |
| n.50G>C | 2 | 121530929 | 30182 | Likely pathogenic | 45 | Y | -0.7194 | UKB/AllofUs | 5' stem loop / k-turn |
| n.51G>A | 2 | 121530930 | 30178 | Pathogenic/Likely pathogenic | 46 | Y | -0.3997 | UKB/AllofUs | 5' stem loop / k-turn |
| n.53C>G | 2 | 121530932 | 30183 | Pathogenic/Likely pathogenic | 48 | N |  |  | 5' stem loop |
| n.55G>A | 2 | 121530934 | 30179 | Pathogenic | 50 | Y | -0.3524 | UKB/AllofUs | 5' stem loop |
| n.111G>A | 2 | 121530990 | 30181 | Pathogenic |  |  |  |  | 3' stem loop |
| n.114G>C | 2 | 121530993 | 977780 | Pathogenic | 116 | Y | -0.1376 | unobserved | 3' stem loop |
| n.116A>C | 2 | 121530995 | 1373455 | Pathogenic | 118 | Y | -0.7121 | UKB/AllofUs | Sm protein binding |
| n.116A>G | 2 | 121530995 | 870579 | Likely pathogenic | 118 | Y | -0.8435 | UKB/AllofUs | Sm protein binding |
| n.116A>T | 2 | 121530995 | 812960 | Likely pathogenic | 118 | Y | -0.0153 | UKB/AllofUs | Sm protein binding |
| n.117A>G | 2 | 121530996 | 1525441 | Pathogenic | 119 | Y | -0.6861 | UKB/AllofUs | Sm protein binding |
| n.120T>G | 2 | 121530999 | 977779 | Pathogenic | 122 | Y | -0.6976 | unobserved | Sm protein binding |
| n.124delG | 2 | 121531002 | 1224468 | Likely pathogenic | 126 | N |  |  | Sm protein binding |
| n.124G>T | 2 | 121531003 | 2813456 | Pathogenic | 126 | N |  |  | Sm protein binding |
| n.124G>A | 2 | 121531003 | 39443 | Pathogenic/Likely pathogenic | 126 | N |  |  | Sm protein binding |
| n.23C>T | 2 | 121530902 | 1133987 | Likely benign |  |  |  |  | loop region |
| n.41G>A | 2 | 121530920 | 1629605 | Likely benign | 36 | N |  |  | loop region |
| n.42C>T | 2 | 121530921 | 1160662 | Likely benign | 37 | N |  |  | loop region |
| n.43A>G | 2 | 121530922 | 1633266 | Likely benign | 38 | N |  |  | loop region |
| n.45A>C | 2 | 121530924 | 1632468 | Likely benign | 40 | N |  |  | loop region |
| n.58C>T | 2 | 121530937 | 1170904 | Benign |  |  |  |  | loop region |
| n.65C>T | 2 | 121530944 | 1631270 | Likely benign |  |  |  |  | Stem I |
| n.87C>T | 2 | 121530966 | 1168718 | Benign/Likely benign |  |  |  |  | 3' stem loop |
| n.93G>A | 2 | 121530972 | 1601357 | Benign |  |  |  |  | 3' stem loop |
| n.109T>A | 2 | 121530988 | 1628512 | Likely benign |  |  |  |  | 3' stem loop |
| n.117dup | 2 | 121530994 | 1159385 | Likely benign |  |  |  |  | Sm protein binding |

**Supplementary Table 6. Oligonucleotide sequences used in this study.** (provided as separate file)

